# Sustainable Development Goals and Health Sector Strategic Indicators Assessment in Cameroon: A Retrospective Analysis at Regional and National Levels

**DOI:** 10.1101/2024.11.25.24317921

**Authors:** Fabrice Zobel Lekeumo Cheuyem, Brian Ngongheh Ajong, Adidja Amani, Lionel Berthold Keubou Boukeng, Christelle Sandrine Ngos, Florence Kissougle Nkongo, Martine Golda Mekouzou Tsafack, Esther Andriane Bitye Bi Mvondo, Guy Stephane Nloga, Ariane Nouko, Michel Franck Edzamba, Denetria Ngati Nyonga, Fernande Murielle Mba Fouda, Yollande Ngo Kam, Christian Mouangue

## Abstract

**Background:** Cameroon has developed a Health Sector Strategy (HSS) that aims at fostering a healthy and productive human capital. To achieve this objective, key health indicators have been defined to track progress towards the goal. This study was conducted to provide evidence on the current situation and assess progress towards achieving the HSS and other key health indicators in the Centre Region and in Cameroon.

**Methods:** Data were collected in April 2023 through a through a comprehensive review of online documents and databases. They were collected retrospectively till 2022. Data were retrieved from the national database using DHIS2 version 2.40 and checked for completeness. Additional data were gathered from online surveys and reports available including Demographic and Health Survey, Multiple Indicators Cluster Survey, National Statistical Institute report for the Centre Region.

**Results:** At national level, the highest performance was observed in the proportion of HIV-positive pregnant women receiving ART (99%), while the lowest performance was observed in the reduction of the prevalence of chronic malnutrition among children under five, with 63% of the target achieved. In the Centre Region, the national target of reducing the prevalence of chronic malnutrition to 11% was achieved. The lowest performance was observed in the proportion of children under five sleeping under long-lasting insecticidal nets. The density of health facilities was above target at national level (2.3 per 10,000 population) but below target at regional level (1.4 per 10,000 population). Coverage of tracer antigens (Penta 3, BCG) decreased over the five-year study period, falling below the national target of 95% in 2022. However, coverage remained above the national average throughout the study period. Coverage of pregnant women attending four or more antenatal clinics and receiving at least three doses of intermittent preventive treatment (IPT) remained low from 2018 to 2022.

**Conclusions:** Significant progress has been made in strengthening the healthcare system and fostering a healthy, productive workforce. Nevertheless, further efforts are required to prevent malaria transmission, promote antenatal care utilization among pregnant women, and address nutritional insecurity in certain Regions.

## Background

Sustainable development refers to the principle of achieving human development goals while maintaining the capacity of natural systems to provide the natural resources and ecosystem services on which economies and societies depend [1]. The third objective of Sustainable Development Goals (SDGs) by 2030 is to ensure the well-being of women and children [2]. Based on these objectives, Cameroon has developed a health sector strategy (HSS) that aims to develop healthy, productive human capital capable of driving strong, inclusive and sustainable growth [3].

One of the key challenges to achieving this goal is malaria, which remains a leading cause of death in sub-Saharan Africa and accounting for 94% of global morbidity [4]. In Cameroon, the national malaria prevention strategy includes promoting the use of long-lasting insecticidal nets (LLINs), intermittent preventive treatment (IPT) for pregnant women and seasonal malaria chemoprevention for children aged 3-59 months in the northern Regions [5].

Moreover, the maternal and neonatal mortality remained a significant concerns in Cameroon [6]. Most maternal deaths are preventable, as effective solutions to address complications are well established. Ensuring that all women have access to quality care during pregnancy, childbirth, and the postpartum period is essential. Maternal and newborn health are intrinsically linked. It is particularly important that all births are attended by skilled healthcare workers, as prompt care and treatment can mean the difference between life and death for both mother and newborn. Additionally, preventing unintended pregnancies is a key aspect of improving maternal health [7].

HIV/AIDS-related deaths have declined, partly due to the scale-up of antiretroviral treatment (ART) [8]. However, women continue to face barriers to ART uptake and adherence, and health systems and structural interventions may have limited capacity to facilitate ART uptake and adherence if they do not also address interpersonal barriers related to partners [9]. Addressing these barriers and improving known facilitators will require the development of respectful and locally acceptable models of HIV service delivery and antenatal care that respond to women’s needs and perspectives, and support them as they enter and progress through the maternal ART cascade [10].

Routine immunization remains a highly cost-effective public health intervention for reducing child mortality [11]. Immunization of children aged 0–11 months and pregnant women is a priority objective in Cameroon’s health strategy [12]. This strategy aims to protect children from infectious diseases, but they still face nutritional problems due to the lack of micronutrients such as iron, deficiency of which remains high in Cameroon, especially among children under 5 and women of childbearing age. Malnutrition in Cameroon has multifactorial causes, including general food insecurity with limited dietary diversity, infrastructural problems such as limited access to basic social services, sanitation infrastructure, and inadequate health care and hygiene practices [13]. There are few reports assessing the achievement of key health indicators in Cameroon. This justifies the conduct of this study, the objective of which was to carry out a situational analysis and assess progress towards the achieving of HSS indicator and health-related SDGs at regional and national levels.

## Methods

### Study Type and Period

Data were collected through a document review and from online data base during April 2023. According to their availability, data were collected retrospectively from till 2022.

### Study Site

Cameroon has an estimated population of 27.6 million in 2022, divided into ten administrative Regions among which the Centre Region. The country has two capitals: Yaounde in the Centre Region, which serves as the political hub, and Douala in the Littoral Region, which drives the country’s economy. Notably, Cameroon’s population is predominantly youthful, with 43.6% of citizens under the age of 15 [14]. The Center Region includes 32 Health Districts, which are urban, semi-urban and rural. This Region is also marked by significant cultural and socio-economic diversity, with numerous ethnic groups contributing to its vibrant cultural landscape and varying economic activities [15] (Figure 1).

**Fig. 1.**
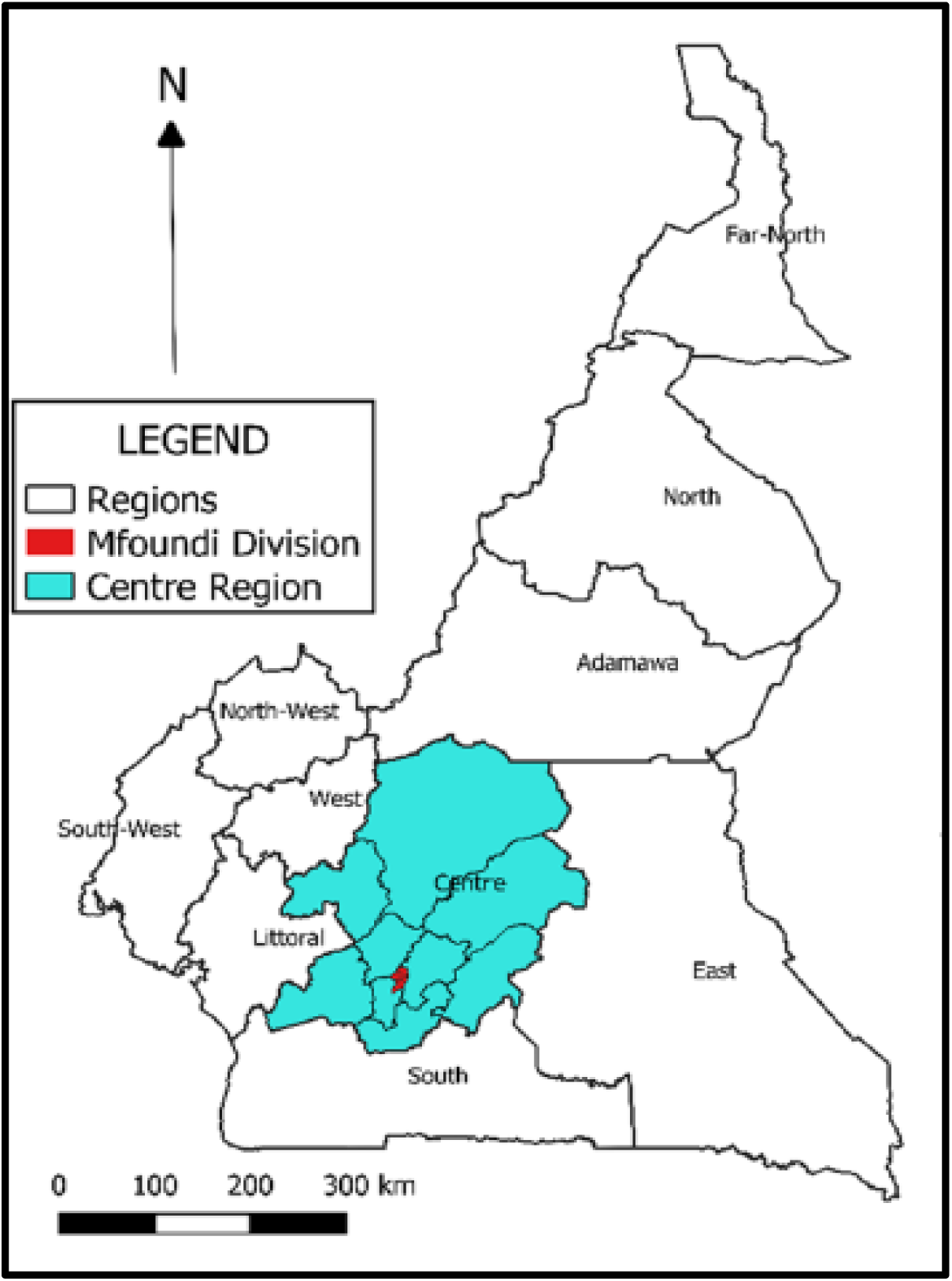
Map of Cameroon highlighting the Centre Region [16].

### Data Sources, Processing and Analysis

Data were retrieved from the online database in using the DHIS2 version 2.40 data visualizer tool, national health data were exported into a Microsoft Office Excel 2019 file. According to national guidelines, this database is updated monthly with reports from health facilities in each health district. To ensure reliability, the extracted the extraction was done by two independent and trained DHIS2 specialists and in case of discrepancy, a third one was called. Data underwent rigorous checks for accuracy and completeness prior to data analysis. Some key indicator data were collected from surveys and reports available online such as Demographic and Health Survey, Multiple Indicators Cluster Survey, National Statistical Institute Report for the Centre Region) [17–19]. Charts presenting various trends were produced using Microsoft Office Excel 2016.

### Variable Selection

Our key indicators were selected from the Global Reference List of 100 core health indicators for outcome monitoring, a standard set of indicators identified by the international community to provide concise information on health status and trends, including responses at the national and international levels. The list includes indicators relevant for reporting at the national, regional or global level and covers all global health priorities related to the Millennium Development Goals agenda [20]. At the national level, the HSS 2020-2030 has identified a set of key indicators, organized into components within a logical framework for monitoring the evolution of the health situation across all levels of the health pyramid [3]. Our analysis focused on the indicators defined in the 2020-2030 framework and was complemented by other key indicators from the WHO global list. The data collected included various components of health systems strengthening, health promotion, disease prevention and case management. Indicators related to the SDGs include vaccination coverage with some tracer antigen (Penta 3, BCG, Measles-Rubella vaccine) and use of antenatal care by pregnant women.

### Operational Definition

Qualified HCWs include Physicians, other clinicians, state-registered nurses, and midwives [21]. Improved sanitation includes a slab pit latrine, a ventilated improved pit latrine, a flush toilet or a composting toilet [22]. Live expectancy refers to the average number of years a person at a given age may be expected to live based on current mortality rates [23].The perioperative mortality rate is the rate of death (from all causes) before hospital discharge among patients who have undergone one or more surgical procedures during their hospital stay. The neonatal mortality rate is the probability that a child born in a given place in a given year or period will die in the first 28 days of life, expressed per 1000 live births. The maternal mortality ratio refers to the annual number of deaths among women, due to causes related to, or exacerbated by pregnancy or its management (excluding accidental or fortuitous causes), occurring during pregnancy or childbirth or in the 42 days following the end of pregnancy, whatever the duration or type of pregnancy, expressed per 1000 live births, over a specified period [6,24].

## Results

## 1. Progress towards Objectives

At the national level, the highest performance was observed in the proportion of HIV-positive pregnant women receiving ART (99%) in 2022, while the lowest performance was observed in the reduction of the prevalence of chronic malnutrition among children under five, with 63% of the target achieved in 2018. On the other hand, in the Centre Region, the national target of reducing the prevalence of chronic malnutrition to 11% was achieved. The lowest performance was observed in the proportion of children under five sleeping under LLINs (Figure 2).

**Fig. 2.**
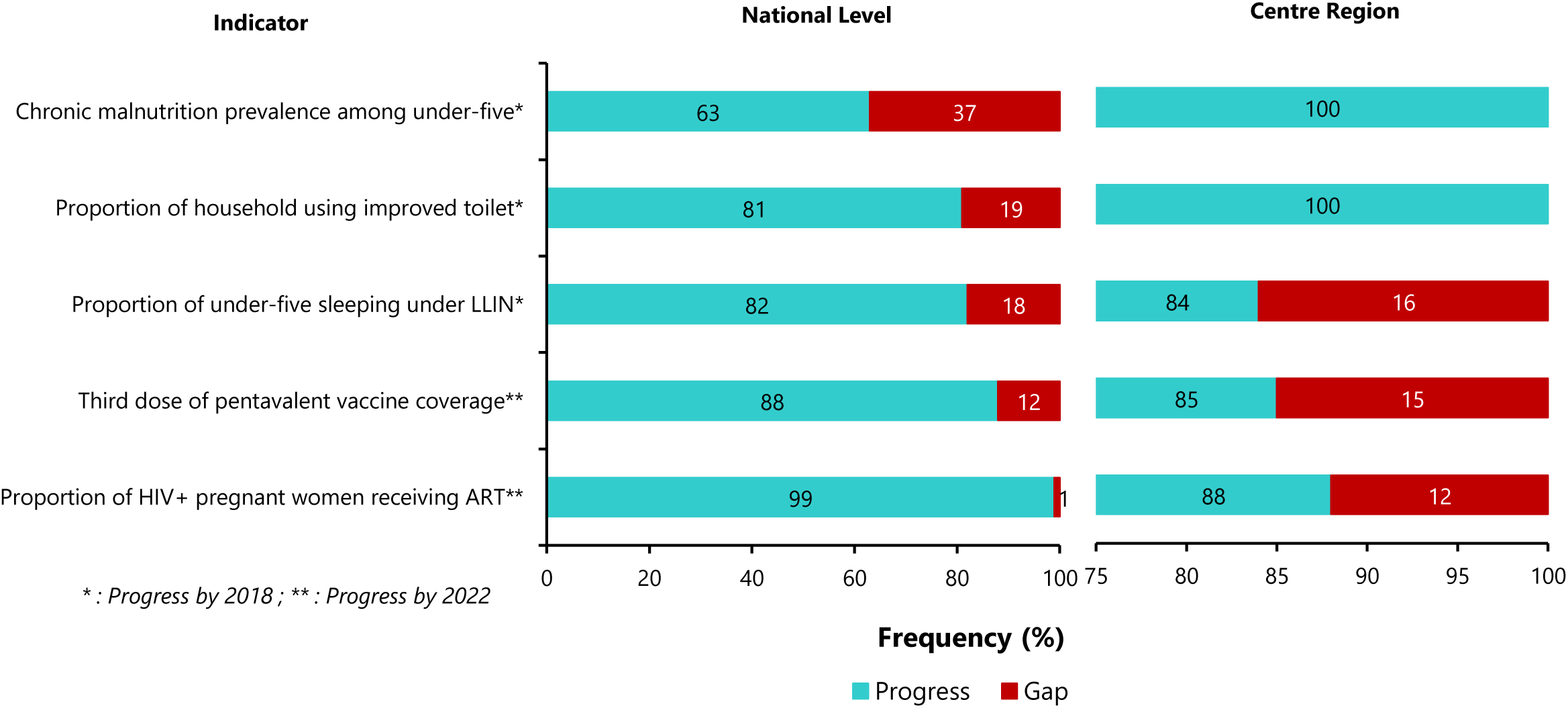
Progress towards achieving some key Health Sector Strategy by 2018 and 2022 at national versus regional level.

## 2. Key indicators of the Health Sector Strategy

### 2.1. Health System Strengthening Component

The density of health facilities was higher than the target at national level (2.3 per 10,000 inhabitants) and lower at regional level (1.4 per 10,000 inhabitants). The density of qualified healthcare workers and the use of services were also below the expected standard at both regional and national levels (Table 1).

**Table 1.** National and Centre Region Indices of availability of health care and services, 2022-2023.

| Indicator | Target | Value |  |
| --- | --- | --- | --- |
| Infrastructure |  | Regional | National |
| Density of health facilities<sup>1</sup> | 2 | 1.4 | 2.3 |
| Hospital bed<sup>1</sup> | 25 | 20.1 | 24.0 |
| Healthcare worker |  |  |  |
| Qualified staff<sup>1</sup> | 23 | 13.7 | 11.3 |
| Use of services |  |  |  |
| Use (outpatient consultations per person/year) | 5 | 0.5 | 0.41 |

<sup>1</sup> Value per 10,000 inhabitants

### 2.2. Impact Indicators

#### 2.2.1. Life Expectancy

Life expectancy at birth has risen sharply over the last decade and will be around 61 years in 2020 (Figure 3).

**Fig. 3.**
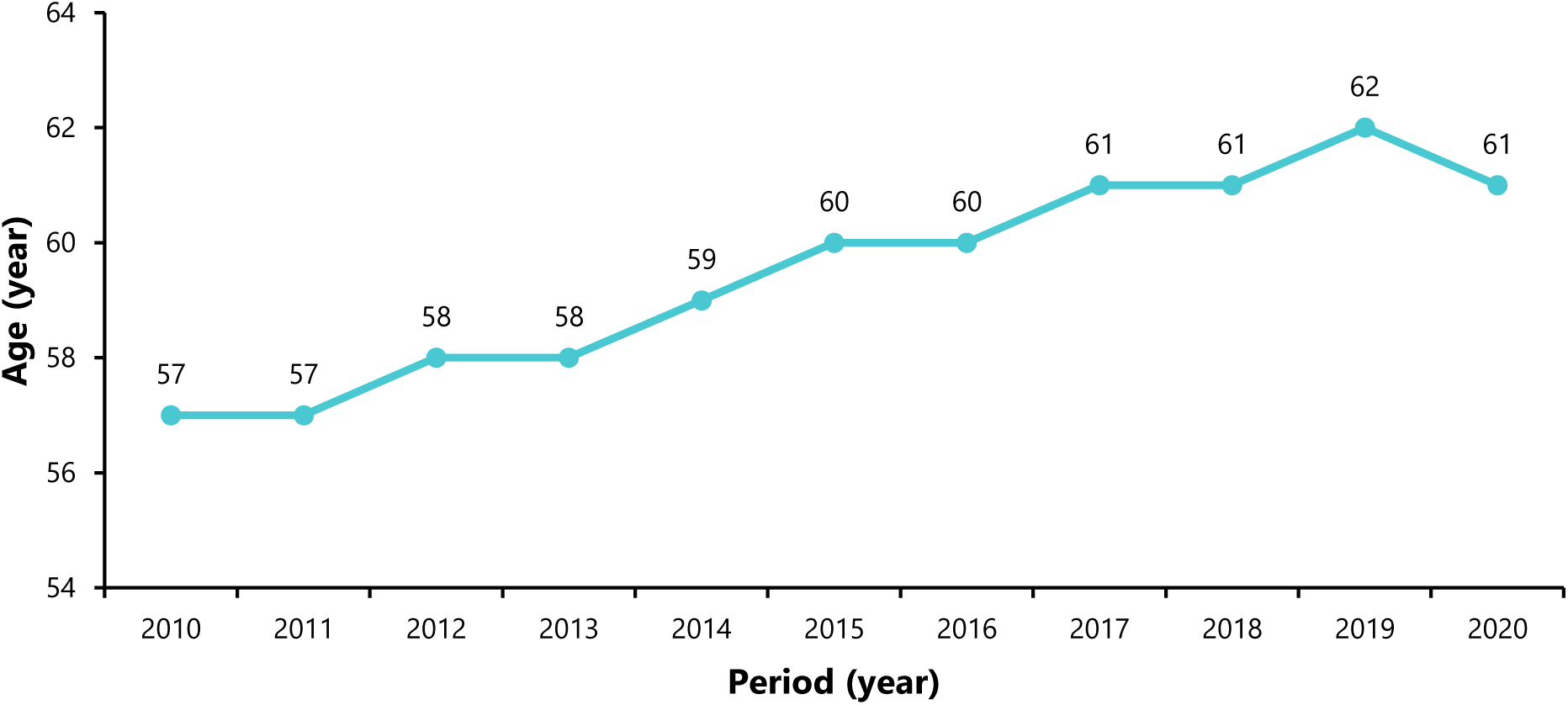
Evolution of life expectancy in Cameroon from 2010 to 2020.

#### 2.2.2. Crude Mortality

The mortality rate in the Centre Region has been increasing since 2019. A peak in mortality was observed in 2021 (3.39 per 1000), before decreasing in 2022. These observations follow the national trends over time (Figure 4).

**Fig. 4.**
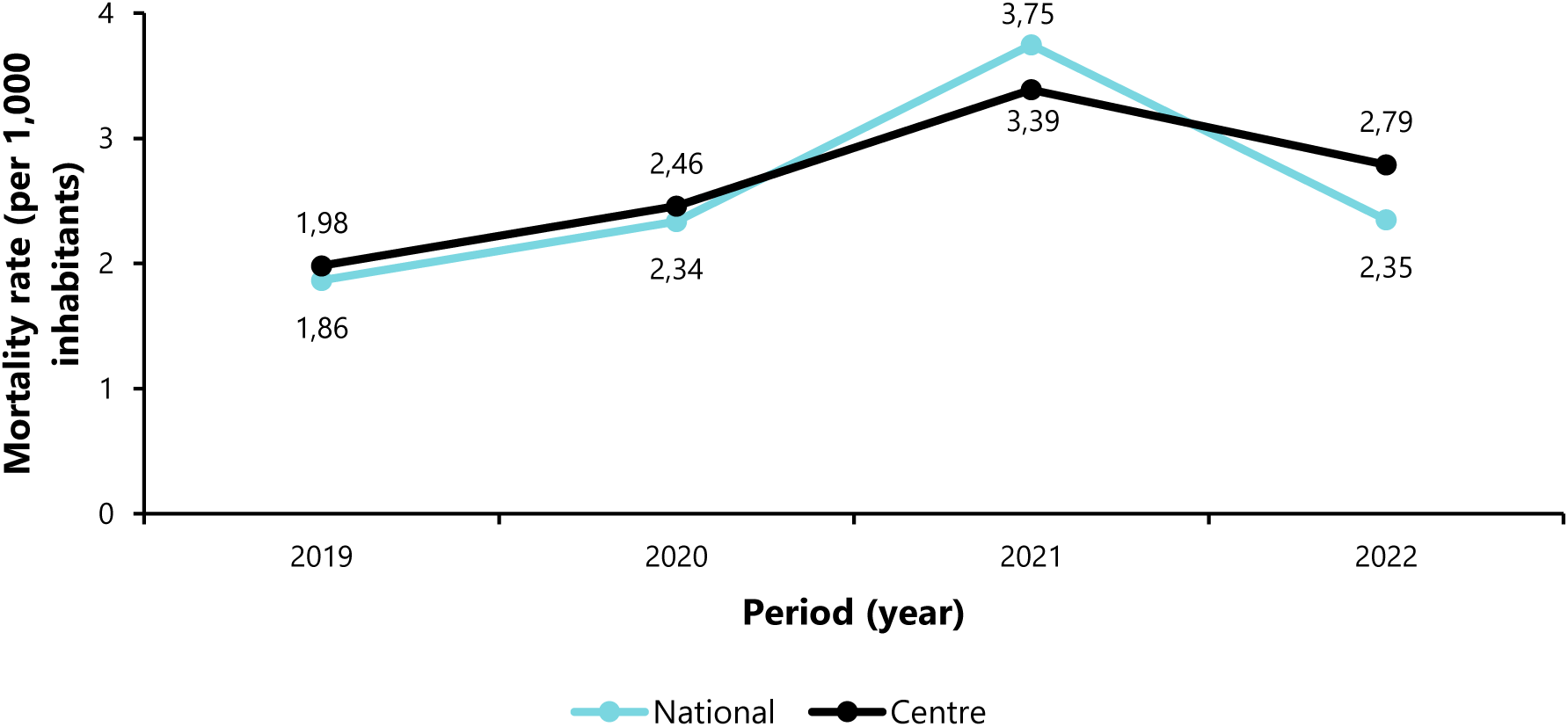
Crude mortality from 2019 to 2022 at national and regional level.

Child-birth related (9.1%) deaths were the leading cause of mortality followed by malaria (7.6%) in the Centre Region. Intestinal diseases, trauma were the third and fourth leading causes of death respectively (Figure 5).

**Fig. 5.**
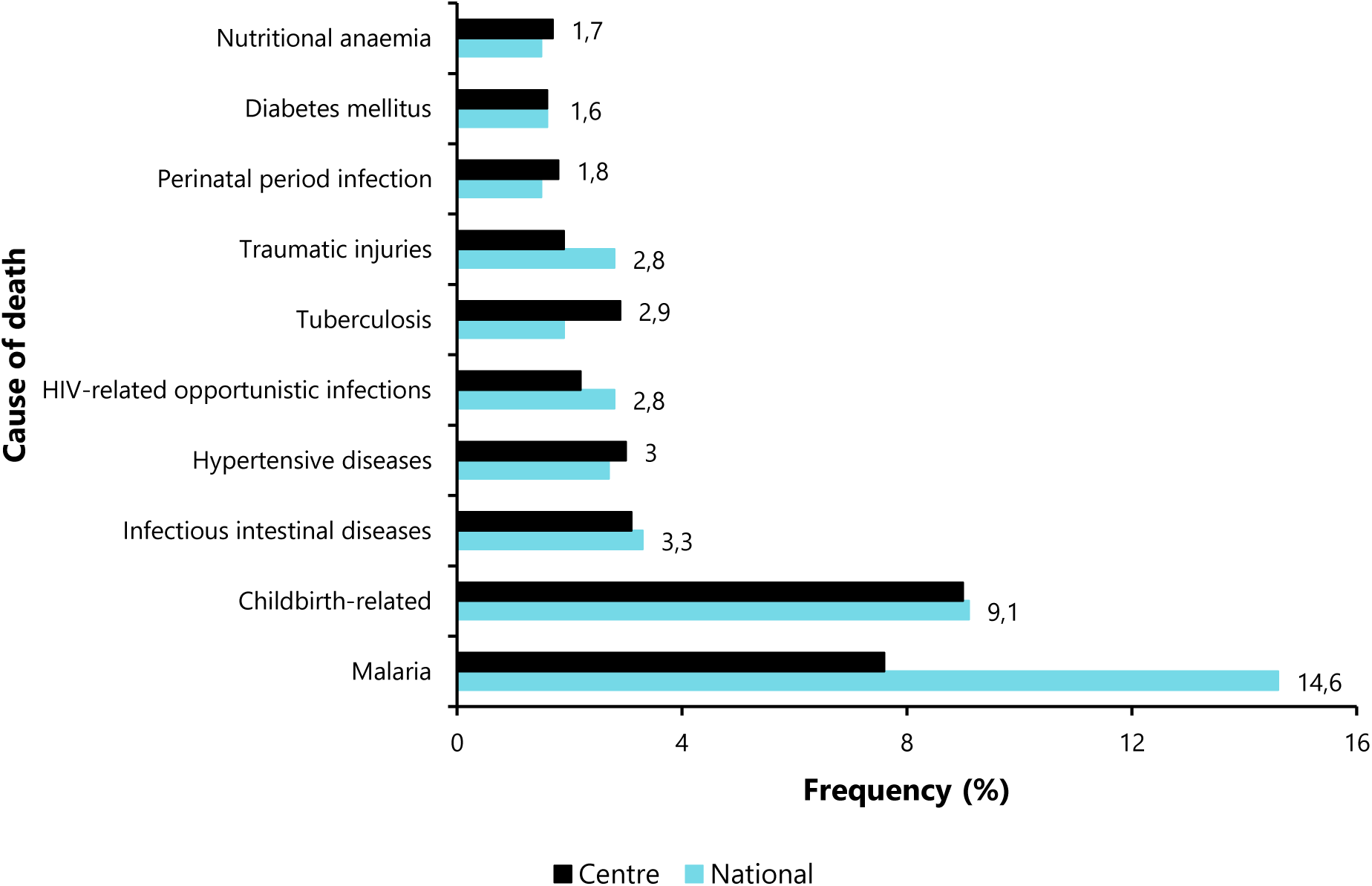
Nine leading causes of death from 2018 to 2022 in the Centre Region and at national level.

### 2.3. Health Promotion Component

#### 2.3.1. Households using Improved Toilets

In the Centre Region, half of households had improved sanitation (56-92%). This trend remained average and stable between 2001 and 2014, and rose sharply to around 90% in 2018. Moreover, these estimates remained higher than the national average, which fluctuated from 40 to 60% over the same period (Figure 6).

**Fig. 6.**
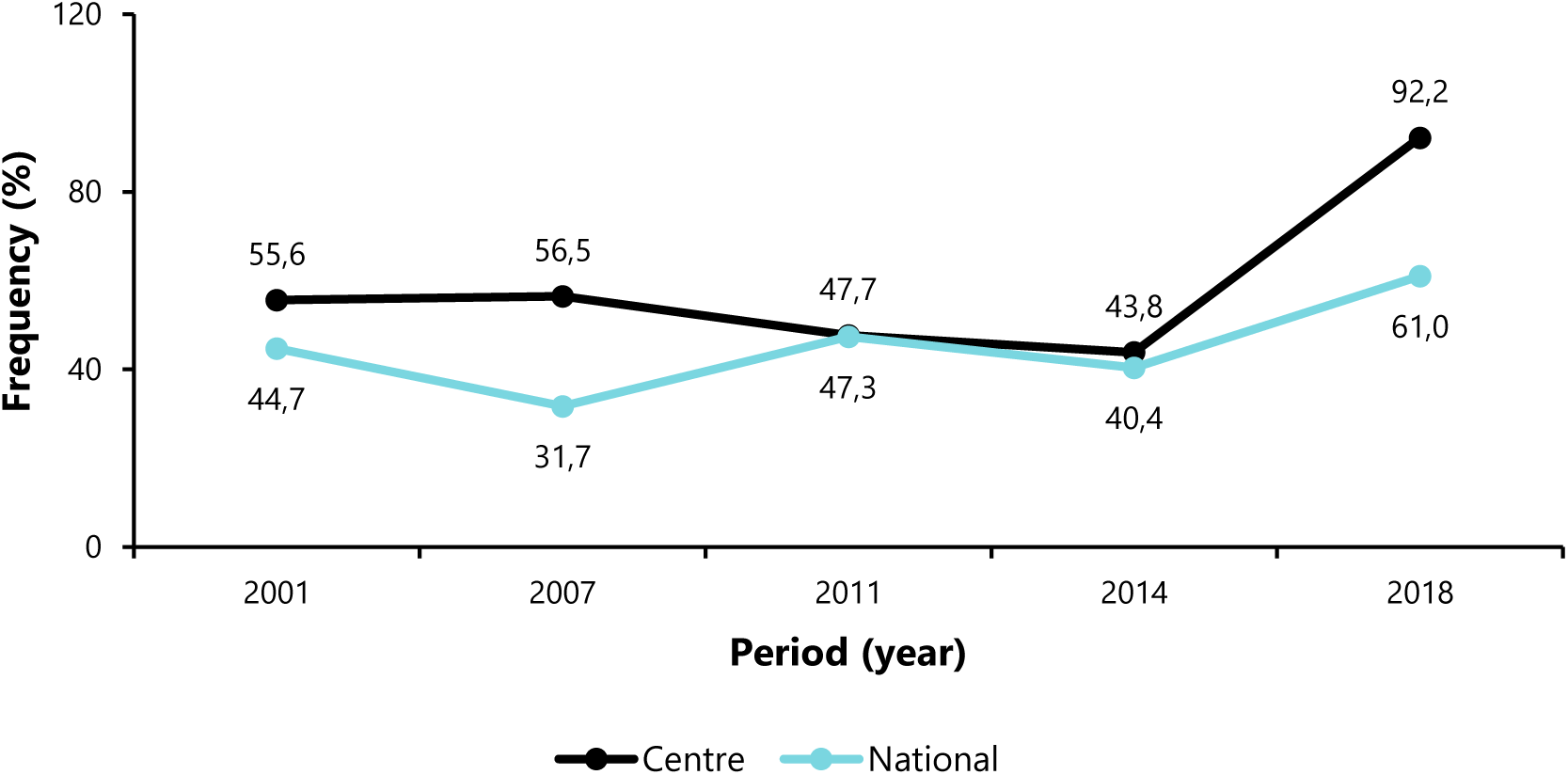
Percentage of households using improved toilets in Centre Region and in Cameroon from 2001 to 2018.

#### 2.3.2. Chronic Malnutrition among Children Under-five

The prevalence rate of this nutritional condition has fallen slightly since 2011 to around 8.9% in 2018 in the Centre Region. This rate remains below the national average (13%) (Figure 7).

**Fig. 7.**
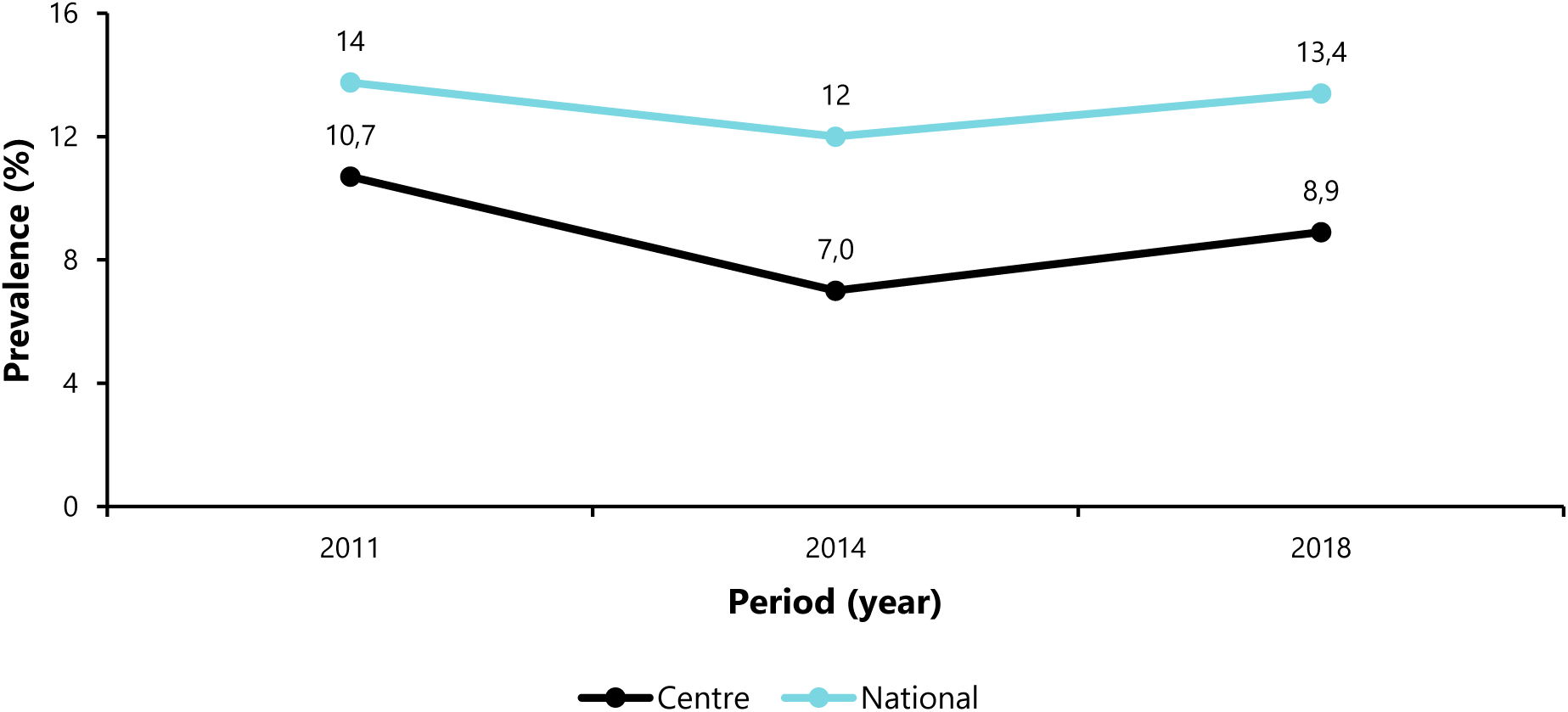
Chronic malnutrition prevalence in children under five from 2011 to 2018 in the Centre Region and in Cameroon.

### 2.4. Disease Prevention Component

#### 2.4.1. Morbidity Burden of Hypertension

In the Centre Region, the prevalence rate increased steadily from 2020 to 2022 at around 3.8%. These estimates remain below the national average (4.7%) (Figure 8).

**Fig. 8.**
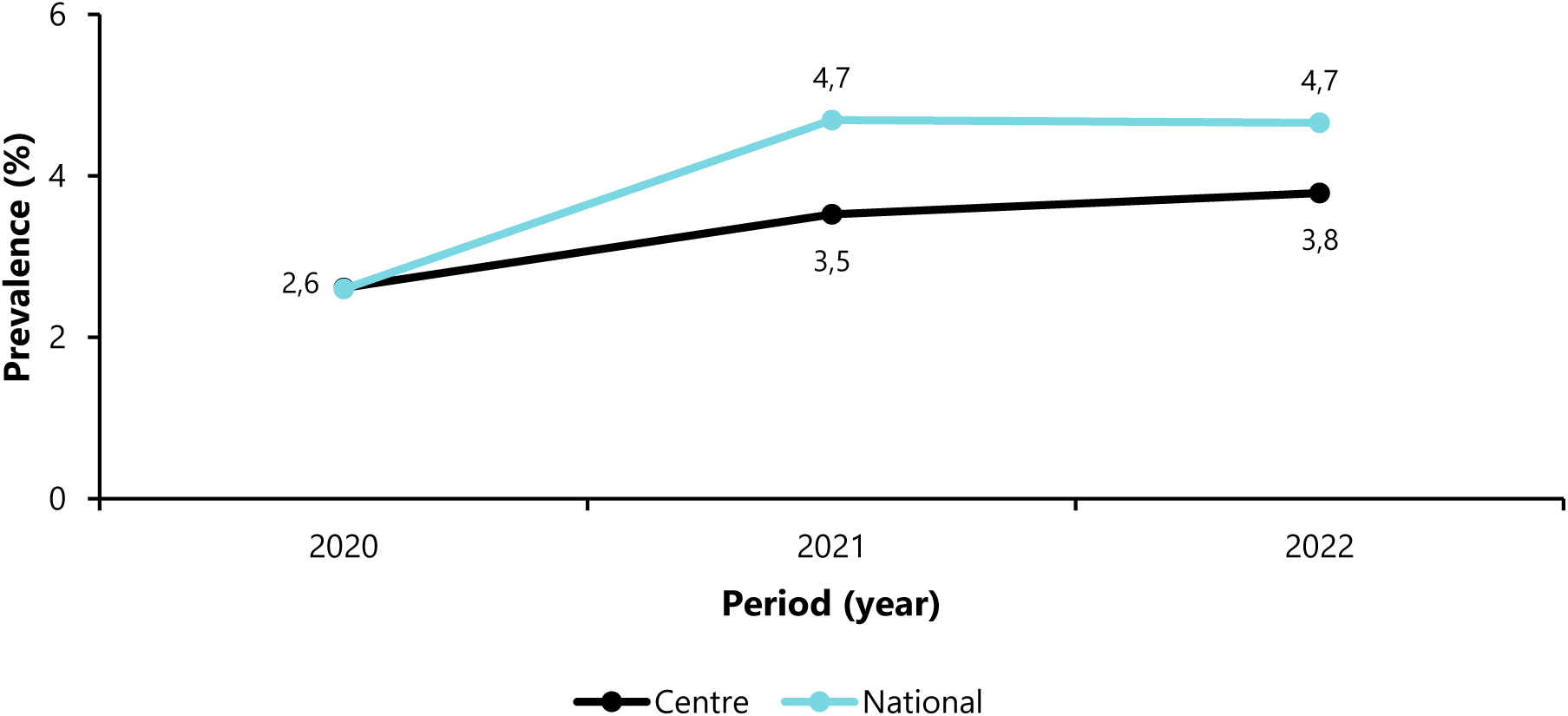
Prevalence of hypertension from 2011 to 2018 in the Centre Region and in Cameroon.

#### 2.4.2. Children Aged 0-5 years Sleeping under Long-Lasting Insecticidal Nets (LLINs)

The trend in the Centre Region shows a significant and steady increase in the proportion of children aged 0-5 sleeping under LLINs, reaching three-quarters of the target in 2018 (75.6%). Estimates followed national trends over the same period (Figure 9).

**Fig. 9.**
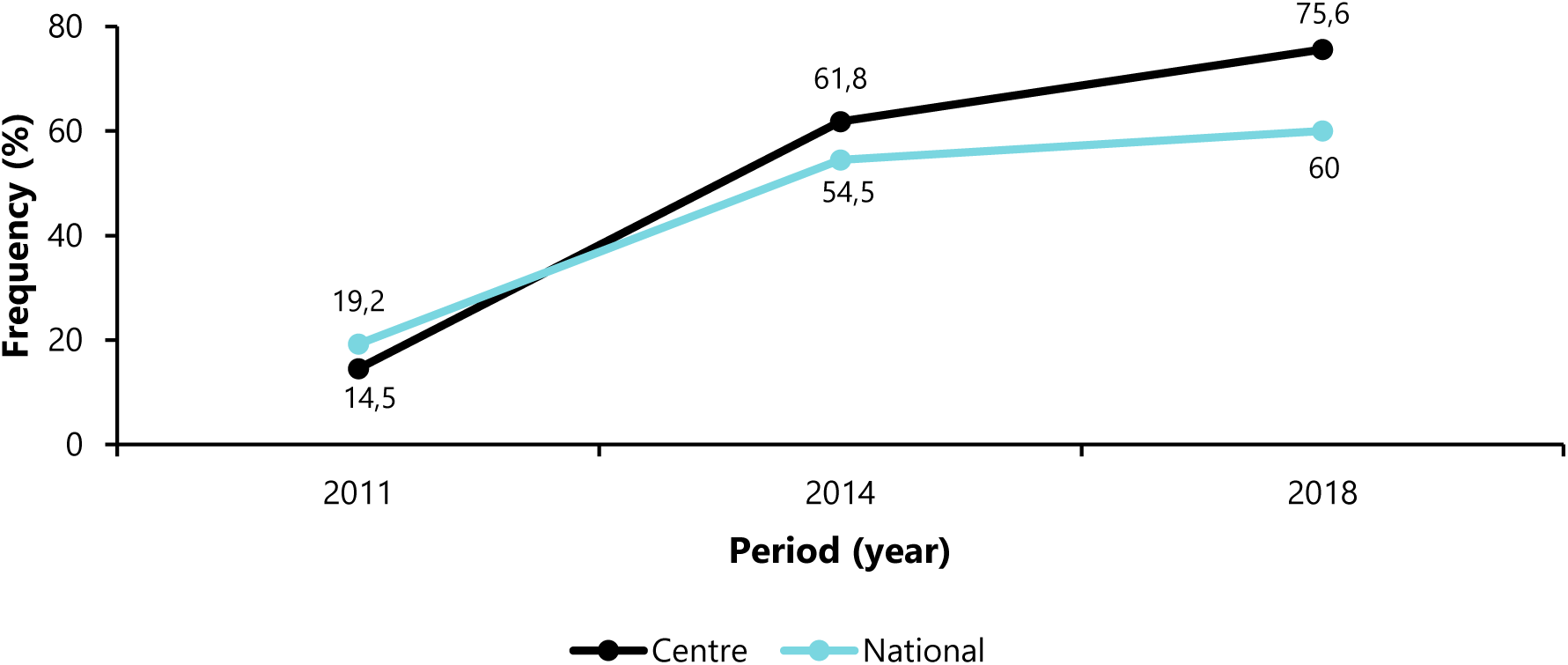
Proportion of children aged 0-5 years sleeping under long-lasting insecticidal nets from 2011 to 2018 in the Centre Region and in Cameroon.

#### 2.4.3. HIV-infected Pregnant Women Receiving Antiretroviral Therapy (ART)

The Centre Region’s performance in enrolling new ART pregnant women has remained below the national average and the 95% target set by the health authorities over the past five years (Figure 10).

**Fig. 10.**
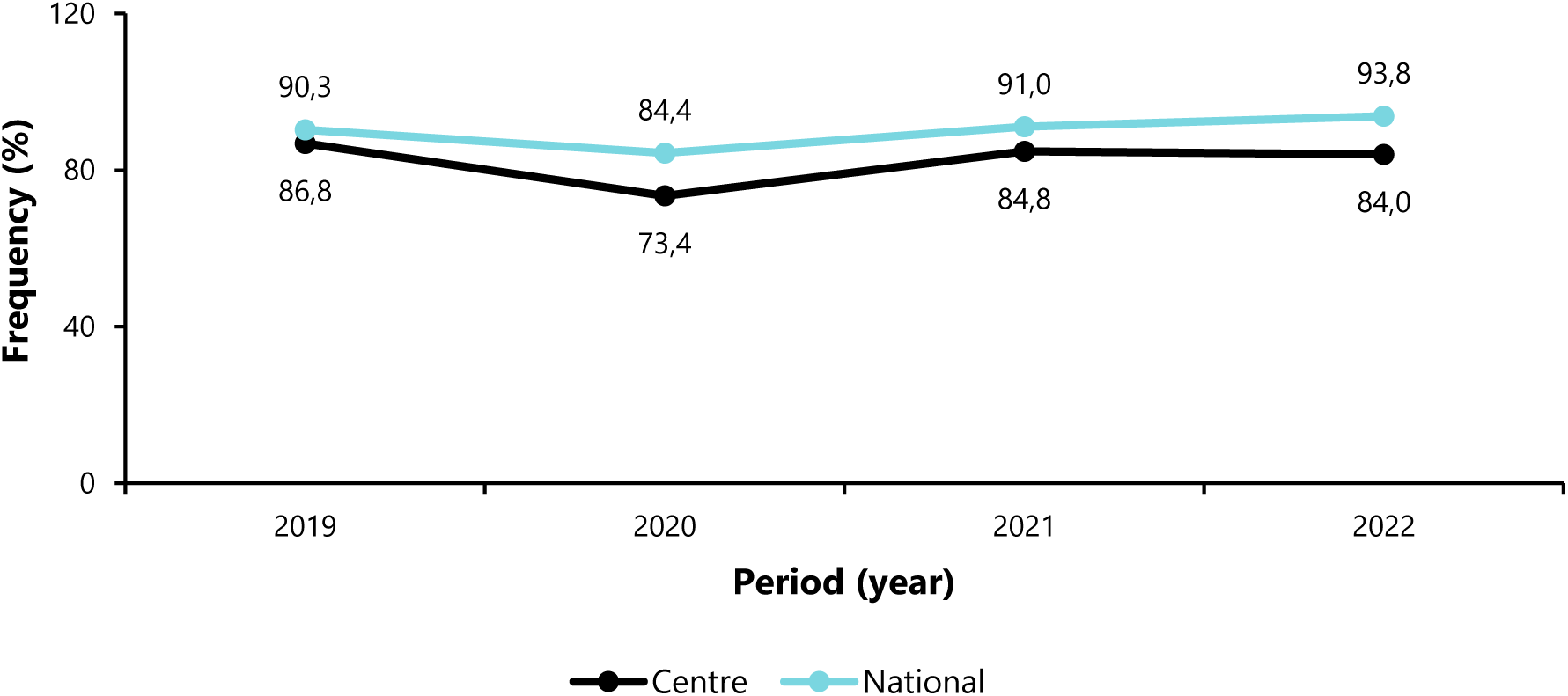
Proportion of newly HIV-infected pregnant women receiving antiretroviral treatment from 2019 to 2022 in the Centre Region and in Cameroon.

### 2.5. Case Management Component

#### 2.5.1. Maternal and Child Mortality

The maternal mortality in the Centre Region has closely mirrored national trends, particularly in 2019 and 2020. However, it has been on a rollercoaster ride, with an upward trend in recent years (2022).

The infant mortality decreased from 43 in 2014 to 39 per 1,000 births in 2018 in the Center Region, reflecting a decrease in mortality among children under one year of age (9.3%).

The neonatal mortality gradually decreases in the Centre Region until 2022 (3.8%) after a slight increase in 2019 (4.9%). Moreover, the regional trends remained higher than those observed at the national level during the same period.

After a peak in infant and child mortality in 2019 (1.7 per 1000 live births), we observe a continuous decrease since then. Moreover, at national level, it has remained above the trends in the Centre Region and stable at around 1.9 per 1000 live births (Figure 11).

**Fig. 11.**
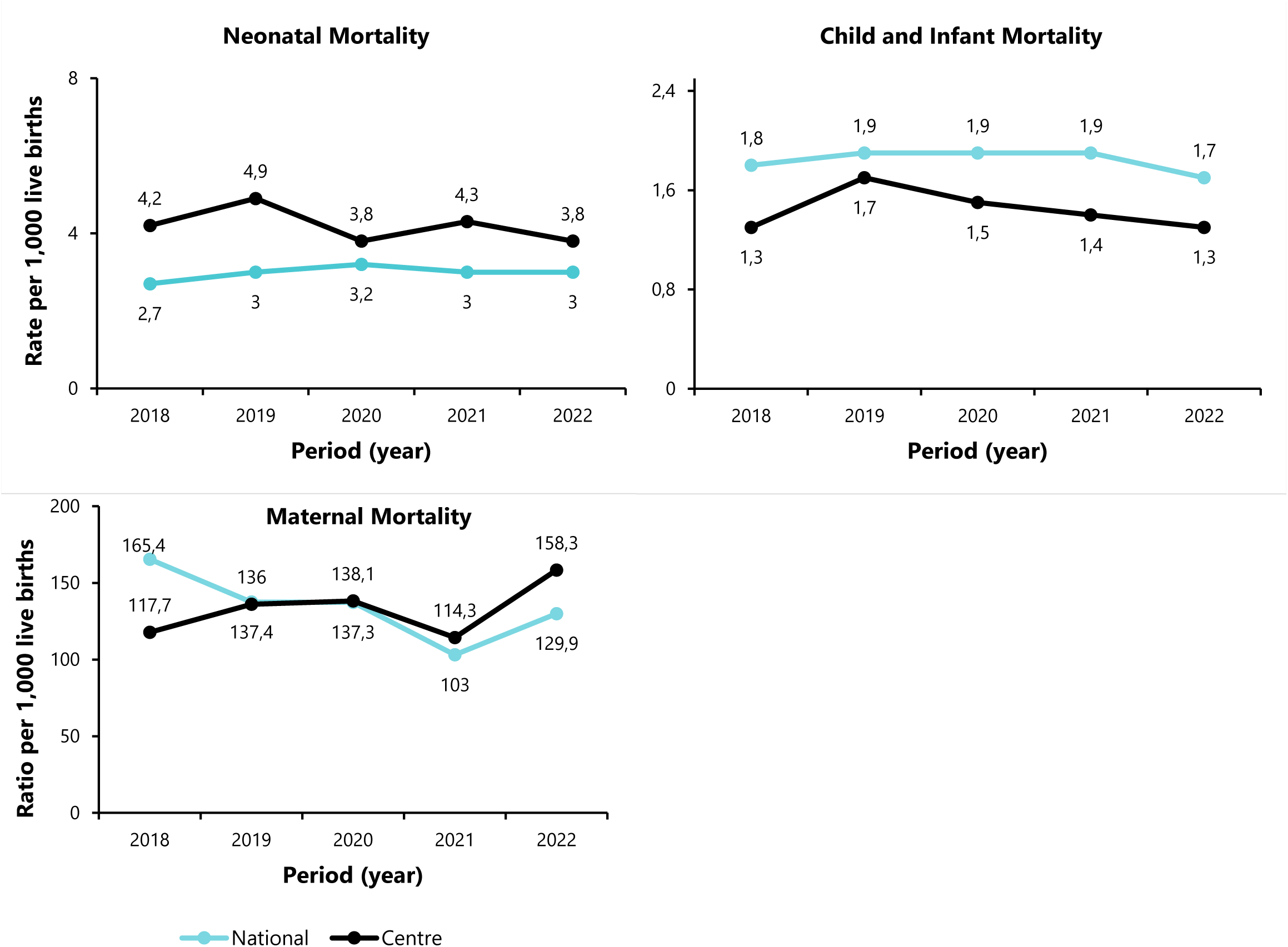
Mortality indicator trends during key period of live among child and women in the Centre Region and in Cameroon.

### Neonatal Mortality

#### 2.5.2. Perioperative Mortality

In the Centre Region, the perioperative mortality was the highest for children under five. However, this rate has decreased over the last three years (Figure 12).

**Fig. 12.**
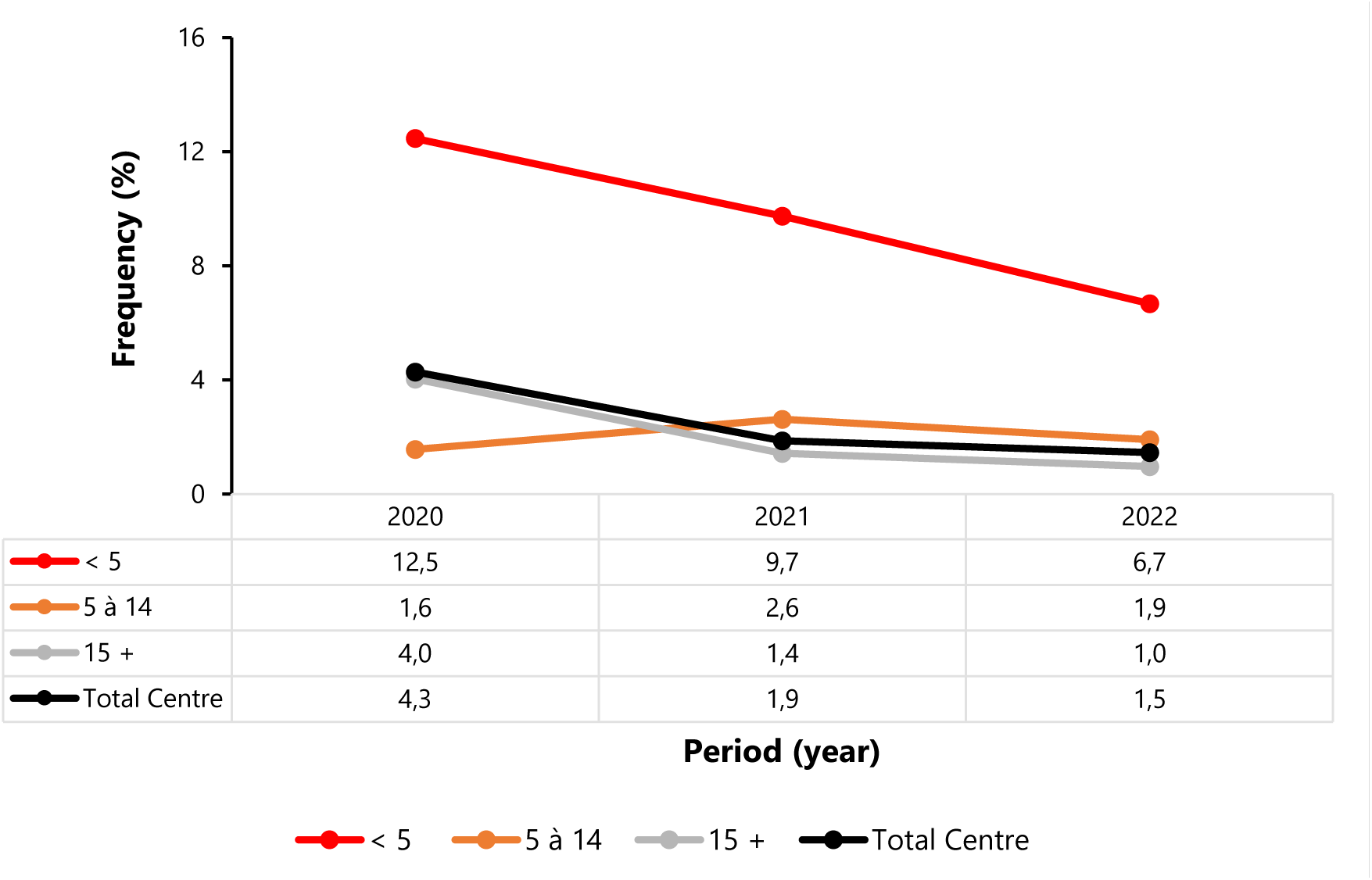
Perioperative mortality by age group from 2020 to 2022 in the Centre Region and in Cameroon.

## 3. SDG Indicator Trend

### 3.1. Vaccination Coverage

In the Centre Region, coverage of tracer antigens (Penta 3, BCG) declined over the five-year study period to below the national target of 90% in 2022. However, coverage remained above the national average throughout the study period (Figure 13).

**Fig. 13.**
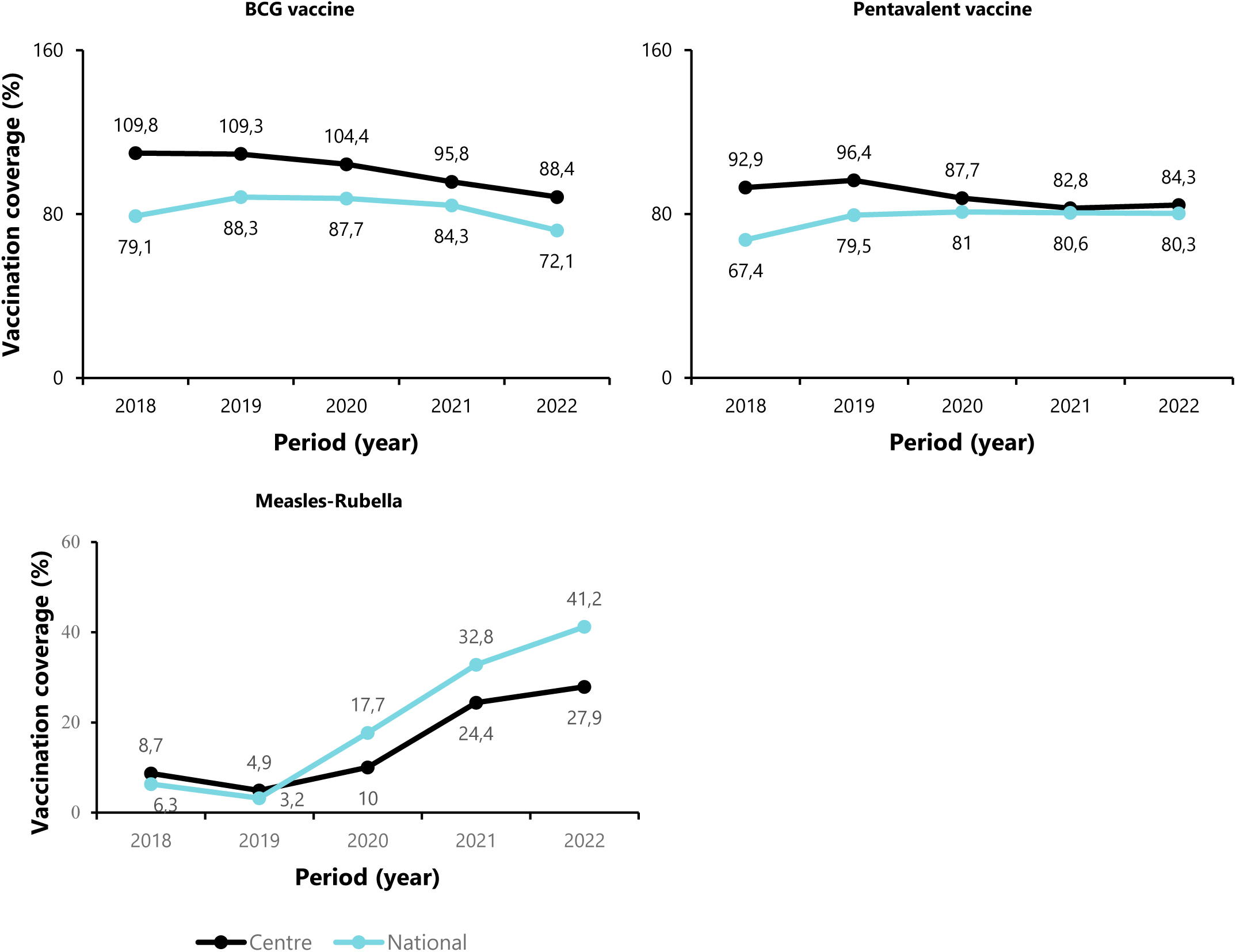
Immunization coverages among 0-11 months from 2018 to 2022 in Centre Region and in Cameroon.

### 3.2. Antenatal Consultation

In the Centre Region, proportion of pregnant women attending four or more antenatal clinics and receiving at least three doses of intermittent preventive treatment (IPT) has remained low from 2018 to 2022. Specifically, IPT3 coverage in 2022 was below the national target of 40-60%. The curves show that coverage in the Centre Region has followed the national average trends while remaining below the national average (Figure 14).

**Fig. 14.**
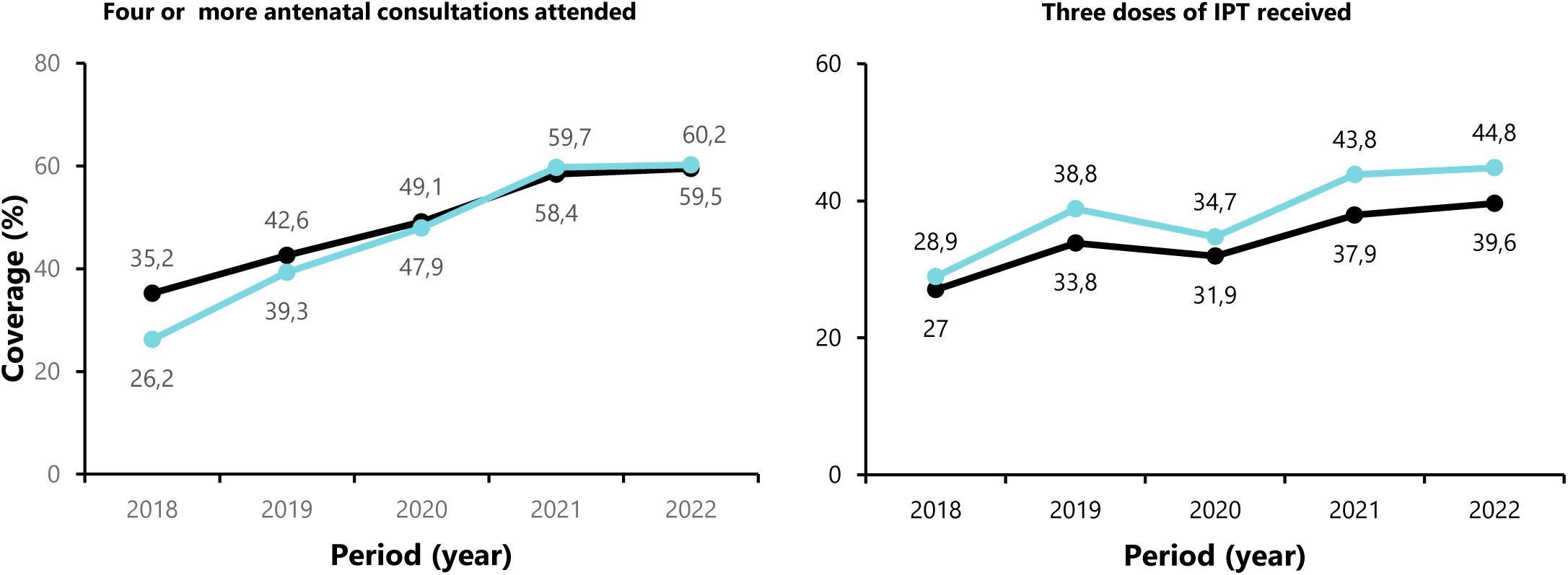
Coverage in Centre Region and Cameroon of pregnant women attending antenatal consultations and receiving an intermittent preventive treatment (IPT) from 2018 to 2022.

## Discussion

### Key Indicator of the Health Sector Strategy

The present study revealed that Cameroon has reached the WHO standard for the availability of health facilities (2.3 per 10,000 population). However, the Centre Region has not yet reached the WHO standard for the availability of health services in all its components [21]. Therefore, considerable efforts are still needed to improve community health services, not only by increasing the density of hospital facilities to bring them closer to the communities, but also by recruiting and ensuring equitable distribution of healthcare workers across the region and area. This will enable HFs to function optimally and improve the attractiveness and utilization of these services by the population.

The mortality trend increases from 2019 and peaks in 2021 before declining in 2022. The deaths due to the COVID-19 pandemic and the related dysfunctions observed, such as the decrease in demand for health services (vaccination, emergency, outpatient consultations) and the quality of health care provided, could explain these trends [25,26]. Similar observations have been made in India [27].

Half of all households in the Central Region had improved sanitation (55-92%) and these proportions were higher than the national average, which varied from 40 to 60% over the same period. This may be related to the fact that community awareness and education on the importance of sanitation and hygiene has led to increased adoption of improved sanitation practices, as well as economic growth and poverty reduction. As economic conditions improve, households are more likely to invest in improved sanitation facilities. Study reports have shown that factors such as zone or location (rural vs. urban), wealth status, and media exposure influence sanitation access and outcomes [28]. These results were higher than those found in the Addis Ababa Region of Ethiopia, where the most of the households did not have improved toilets (92-100%) [29].

The prevalence of chronic malnutrition among under-five children condition has decreased slightly since 2011, to approximately 8.9% in 2018 in the Centre Region and remained lower than the national average (13%). Economic access to nutritious food is an important underlying driver of nutritional outcomes in Cameroon. According to the 2021 Fill the Nutrient Gap in Cameroon study, 48% of households cannot afford a nutritious diet. Infant and young child feeding (IYCF) practices are largely inadequate in the country and can also be considered a major cause of malnutrition) [13]. The rate of exclusive breastfeeding is also low, at only 40% (EDS 2018). In addition to the above factors, a study in Cameroon found that inadequate drinking water consumption was a potentially modifiable risk factor for undernutrition [30].

Regarding the proportion of children under five sleeping on the trend shows a significant and steady increase, reaching three-quarters of the target in 2018 (75.6%). Estimates followed national trends over the same period. The Ministry of Public Health conducted four national mass distribution campaigns of LLINs, the last of which took place in April 2022 [31,32]. This may explain the improvement in household LLIN use, particularly among children under five.

The Centre Region’s performance in enrolling new HIV+ women on ART has remained below the national average and the 95% target set by the health authorities over the past five years. Recent studies have shown that the main barriers to getting pregnant women on treatment include fear of disclosing their HIV status to partners and family members, stigma and discrimination. Knowing their HIV status before becoming pregnant is one of the key drivers of adherence among women on ART [33].

Infant mortality fell from 43 per 1,000 births in 2014 to 39 per 1,000 births in 2018 in the Centre Region, reflecting a drop in mortality among children under one year of age (9.3%). Measures to improve child nutrition, prevent disease (immunization, LLINs), control epidemics (measles, tetanus, yellow fever) and bring health facilities closer to the population may explain this trend. Similar interventions elsewhere have led to satisfactory reductions in child mortality [34].

The neonatal mortality gradually decreases in the Centre Region until 2022 (3.8%) after a slight increase in 2019 (4.9%). Moreover, the regional trends remained higher than those observed at the national level during the same period. To reduce neonatal mortality, it is necessary to improve working conditions (by increasing the number of staff and equipment) and to train and retrain health workers in emergency obstetric and neonatal care. More specifically, the strategy for tackling this problem is based on the implementation of a policy of caring for newborns, especially those with complications of premature birth, birth asphyxia and infections (laboratory tests), before the parents pay for care. This significantly reduces waiting times and improves newborn survival rates [35,36].

The infant and child mortality rate in the Centre Region remained stable at around 1.9 per 1,000 live births from 2018 to 2022. In Kenya, demographic and health surveys show higher rates (6-12%). Premature birth, pneumonia, congenital malformations, neonatal infections, malaria, sepsis, measles, complications of childbirth and diarrhea are the leading causes of preventable death in children under five [37,38].

According to relevant guidelines on good nutrition (immediate and exclusive breastfeeding, access to food and micronutrients) is fundamental to preventing many deaths. In addition to this general measure, access to skilled care for antenatal, delivery and postnatal care, family knowledge of the signs of danger to a child’s health, improved access to water, sanitation, hygiene and immunization all help to prevent child deaths [37,38].

Perioperative mortality is a key measure of the quality of hospital care. In the Centre Region, the peri-operative mortality rate was highest for children under five years of age. However, this rate has decreased over the last three years. As the under-fives are a vulnerable group, this may explain their poorer post-operative prognosis. A study in Nigeria found that the oldest age group was most affected [39]. Improving the quality of care in reception, anesthetic consultation, technical support and post-operative care are levers that would enable the health facility to improve this indicator [40].

### SDG Indicator – Vaccination and Antenatal Consultation Coverage

Vaccination coverage in the Center Region for tracer antigens (Penta 3, BCG) has declined over the past five years and is now below the national target of 95%. However, coverage remained above the national average during the studied period. In the aftermath of the COVID-19 crisis, vaccination services experienced a significant drop in attendance due to fears of contamination, all in the context of the containment ordered by the administrative authorities in March 2020.

Globally, the COVID-19 pandemic had a negative impact on routine childhood immunization coverage and immunization services worldwide. In developing countries such as Cameroon, where immunization coverage was already lower than in developed countries, the impact of the pandemic was even more pronounced, increasing the likelihood of outbreaks of vaccine-preventable diseases and widening existing inequalities. The implementation of catch-up activities in all settings to maintain vaccination coverage is essential to protect vulnerable populations and avert further health crises [41].

In the Central Region, coverage of pregnant women attending four or more antenatal care visits and receiving at least three doses of intermittent preventive treatment (IPT) has remained low over the past five years. Specifically, IPT 3 coverage in 2022 was below the national target of 40-60% [42]. The reasons for this poor performance were assessed, and the main factors associated with low attendance at health facilities by women of childbearing age in both rural and urban areas were identified as lack of decision-making autonomy, geographical inaccessibility, financial inaccessibility, self-medication, reliance on traditional practitioners, poor quality of services (poor reception, negligence, stock-outs of drugs).

### Limitations

This study relied on secondary data, which may have limitations in terms of data collection and transmission quality. However, the National Health Survey data utilized in this study were collected using a rigorous methodology, minimizing the risk of bias. Nevertheless, given that the most recent data available are from 2018 in Cameroon, there is a need to conduct a new national health survey to generate more up-to-date and accurate data for decision-making.

### Conclusions

Based on the key performance indicator assessed in this study, significant progress has been made in improving the health system and developing healthy, productive human capital. However, efforts are still needed to reduce nutritional insecurity in some regions of the country. For the Central Region in particular, resources should be directed towards promoting the use of LLINs for malaria prevention and the use of antenatal care by pregnant women to ensure safe pregnancy and delivery.

## Declarations

### Author contributions

Study design & conception: FZLC; Data collection: FZLC, CM; Data analysis, visualization and interpretation: FZLC; Drafting of original manuscript: FZLC; Critical revision of the manuscript: FZLC, BNA, AA, LBKB, CSN, FKN, MGTM, EABBM, GSN, AN, MFE, DNN, FMMF, YNG, CM. final approval of the manuscript: All authors.

### Ethical Approval Statement

Ethical clearance for the present study was waived by the Faculty of Medicine of Yaounde ethical review board. As this secondary data was anonymous and publicly available in aggregated data, there was no risk to the individuals whose data was collected. All methods were performed in accordance with the relevant guidelines of the Helsinki declaration.

### Consent for publication

Not applicable.

### Availability of data and materials

All data generated or analyzed during this study are included in this published article:

## Data Availability

All data produced in the present work are contained in the manuscript

## Abbreviations

BCG: Bacillus Calmet Guerin
COVID-19: New Coronavirus Disease
HIV: Human Immunodeficiency Virus
HSS: Health Sector Strategy
DHIS: District Health Information Software
AIDS: Acquired Immunodeficiency Syndrome
ART: Antiretroviral Therapy
Penta: Pentavalent Vaccine
SDG: Sustainable Development Goal
LLIN: Long-Lasting Insecticidal Net
IPT: Intermittent Preventive Treatment

## Reference

1. Mensah J. Sustainable development: Meaning, history, principles, pillars, and implications for human action: Literature review. Ricart Casadevall S, editor. Cogent Soc Sci. 2019;5(1):1653531.

2. Objectif 3: Good health and well-being. Joint SDG Fund, New York. 2022. https://jointsdgfund.org/fr/sustainable-development-goals/goal-3-good-health-and-well-being. Accessed:2024 Nov 6.

3. Cameroun - Stratégie sectorielle de santé 2016-2027. Ministry of Public Health, Yaoundé. 2015. https://www.ilo.org/dyn/natlex/natlex4.detail?p_isn=111170&p_lang=fr. Accessed: 2023 Mar 27.

4. Li J, Docile HJ, Fisher D, Pronyuk K, Zhao L. Current Status of Malaria Control and Elimination in Africa: Epidemiology, Diagnosis, Treatment, Progress and Challenges. J Epidemiol Glob Health. 2024;14(3):561–79.

5. Talipouo A, Ngadjeu CS, Doumbe-Belisse P, Djamouko-Djonkam L, Sonhafouo-Chiana N, Kopya E, et al. Malaria prevention in the city of Yaoundé: knowledge and practices of urban dwellers. Malar J. 2019;18(1):167.

6. Cheuyem FZL, Amani A, Ajong BN, Boukeng LBK, Mouangue C, Tsafack MGM, et al. Humanization of Care: A Geospatial Analysis of Key Indicators of Quality and Safety of Health Care and Service in the Centre Region, Cameroon. medRxiv.2024. doi:10.1101/2024.11.11.24317125v1

7. SDG Target 3.1 Maternal mortality. WHO, Geneva. https://www.who.int/data/gho/data/themes/topics/sdg-target-3-1-maternal-mortality. Accessed:2023 Apr 28.

8. HIV and AIDS Epidemic Global Statistics. HIV.gov. 2024. https://www.hiv.gov/hiv-basics/overview/data-and-trends/global-statistics. Accessed: 2024 Nov 21.

9. Omonaiye O, Kusljic S, Nicholson P, Manias E. Medication adherence in pregnant women with human immunodeficiency virus receiving antiretroviral therapy in sub-Saharan Africa: a systematic review. BMC Public Health. 2018;18(1):805.

10. Hodgson I, Plummer ML, Konopka SN, Colvin CJ, Jonas E, Albertini J, et al. A Systematic Review of Individual and Contextual Factors Affecting ART Initiation, Adherence, and Retention for HIV-Infected Pregnant and Postpartum Women. PLOS ONE. 2014;9(11):e111421.

11. Zhou F. Health and Economic Benefits of Routine Childhood Immunizations in the Era of the Vaccines for Children Program — United States, 1994–2023. MMWR Morb Mortal Wkly Rep. 2024; 73:682–5.

12. Amani A, Saidu Y, Asaah CT, Cheuyem FZL, Njoh AA, Kabir HA, et al. Immunization in Cameroon: Uncovering Progress, Confronting Challenges, and Paving the Path towards Achieving Sustainable Development Goal 2030 Targets. *J Vaccines Vaccin.* 2023;14(3):1–10.

13. WFP Cameroon Malnutrition Prevention and Treatment, October 2022 - Cameroon | ReliefWeb. 2022. https://reliefweb.int/report/cameroon/wfp-cameroon-malnutrition-prevention-and-treatment-october-2022. Accessed:2023 Apr 27.

14. Amani A, Nansseu JR, Ndeffo GF, Njoh AA, Cheuyem FZL, Libite PR, et al. Stillbirths in Cameroon: an analysis of the 1998-2011 demographic and health surveys. BMC Pregnancy Childbirth. 2022;22(1):736.

15. Cheuyem FZL, Amani A, Ajong BN, Otsali RKN, Nouko A, Guissana EO, et al. Assessment of Vaccine managers’ Knowledge and Observance of Vaccination Norms and Standards in an Urban Health District of Yaounde (Cameroon) during the COVID-19 Pandemic. medRxiv; 2024 Accessed:2024 Nov 18]. p. 2024.11.07.24316922. https://www.medrxiv.org/content/10.1101/2024.11.07.24316922v3

16. Analyse géographique de la couverture sanitaire au Cameroun Répartition des formations sanitaires et de la charge du personnel de santé. Geomatic Strategy. 2019. https://www.geostrategies.net/2019/04/01/strategies-spatiales-n2-mars-avril-2019-analyse-geographique-de-la-couverture-sanitaire-au-cameroun-repartition-des-formations-sanitaires-et-de-la-charge-du-personnel-de-sante/. Accessed: 2021 Sep 11.

17. Résultat MICS 2014, Cameroun. UNICEF. 2015. https://mics.unicef.org/download-tracker/5396/9936. Accessed: 2023 Apr 13.

18. Enquête Démographique et de Santé 2018. Institut National de la Statistique, Yaoundé. 2019. https://ins-cameroun.cm/wp-content/uploads/2020/06/VIH-EDSC-V.pdf. Accessed: 2023 Apr 27.

19. Annuaire Statistique de la Région du Centre, édition 2019 – Institut National de la Statistique du Cameroun. https://ins-cameroun.cm/en/statistique/annuaire-statistique-de-la-region-du-centre-edition-2019/. Accessed:2023 Apr 27.

20. Liste mondiale de référence des 100 indicateurs sanitaires de base. WHO, Geneva. 2015. https://iris.who.int/handle/10665/204687. Accessed:2023 Apr 9.

21. Organisation mondiale de la Santé. Mesurer la disponibilité et la capacité opérationnelle des services (SARA): un outil d’évaluation des établissements de santé: manuel de référence. Organisation mondiale de la Santé. 2014. https://apps.who.int/iris/handle/10665/149026 Accessed:2023 Jun 12.

22. Installations sanitaires de base. UNESCO. 2020. https://uis.unesco.org/fr/glossary-term/installations-sanitaires-de-base Accessed:2023 Apr 27.

23. Tulchinsky TH, Varavikova EA. Chapter 3 - Measuring, Monitoring, and Evaluating the Health of a Population. In: Tulchinsky TH, Varavikova EA, editors. The New Public Health (Third Edition). San Diego: Academic Press; 2014 p. 91–147.

24. 2018 Global reference list of 100 core health indicators (plus health-related SDGs). WHO, Geneva. 2018. https://www.who.int/publications/i/item/2018-global-reference-list-of-100-core-health-indicators-(-plus-health-related-sdgs). Accessed:2024 Nov 7.

25. Moynihan R, Sanders S, Michaleff ZA, Scott AM, Clark J, To EJ, et al. Impact of COVID-19 pandemic on utilisation of healthcare services: a systematic review. BMJ Open. 2021;11(3): e045343.

26. Mewoabi S. The Impact of Covid-19 Pandemic and Gender-Based Violence on Uptake of HIV Services in Touboro District Hospital, Cameroon. TEXILA Int J PUBLIC Health. 2022;10(1):296–306.

27. K AKA, Mishra N. Mortality during the COVID-19 pandemic: the blind spots in statistics. Lancet Infect Dis. 2022;22(4):428–9.

28. Demsash AW, Tegegne MD, Wubante SM, Walle AD, Donacho DO, Senishaw AF, et al. Spatial and multilevel analysis of sanitation service access and related factors among households in Ethiopia: Using 2019 Ethiopian national dataset. PLOS Glob Public Health. 2023;3(4):e0001752.

29. Belay DG, Andualem Z. Limited access to improved drinking water, unimproved drinking water, and toilet facilities among households in Ethiopia: Spatial and mixed effect analysis. PLOS ONE. 2022;17(4): e0266555.

30. Ngassa AB, Meriki HD, Mbanga CM, Nzefa LD, Mbhenyane X, Tambe AB. Key predictors of undernutrition among children 6–59 months in the Buea Health District of the Southwest region of Cameroon: a cross sectional community-based survey. BMC Nutr. 2022;8(1):148.

31. La 4e campagne nationale de distribution des Milda sera lancée en avril 2022. Atangana O. Cameroun. 2021. https://lurgentiste.com/cameroun-la-4e-campagne-nationale-de-distribution-des-milda-sera-lancee-en-avril-2022/. Accessed:2023 Apr 27.

32. 2eme Campagne nationale de distribution gratuite de 12,350 million de MILDA. MINSANTE, Yaoundé. https://www.minsante.cm/site/?q=fr/content/2eme-campagne-nationale-de-distribution-gratuite-de-12350-millions-de-milda. Accessed:2023 Apr 27.

33. Kanguya T, Koyuncu A, Sharma A, Kusanathan T, Mubanga M, Chi BH, et al. Identifying barriers to ART initiation and adherence: An exploratory qualitative study on PMTCT in Zambia. PLoS ONE. 2022;17(1): e0262392.

34. Karungula J. Measures to reduce the infant mortality rate in Tanzania. Int J Nurs Stud. 1992;29(2):113–7.

35. Mah Mungyeh E, Chiabi A, Tchokoteu Pouasse F, Nguefack S, Bogne J, Siyou H, et al. Neonatal mortality in a referral hospital in Cameroon over a seven-year period: trends, associated factors and causes. Afr Health Sci. 2014;14(3):517–25.

36. Ndombo PK, Ekei QM, Tochie JN, Temgoua MN, Angong FTE, Ntock FN, et al. A cohort analysis of neonatal hospital mortality rate and predictors of neonatal mortality in a sub-urban hospital of Cameroon. Ital J Pediatr. 2017;43(1):52.

37. Child mortality (under 5 years). WHO, Geneva. 2020. https://www.who.int/news-room/fact-sheets/detail/levels-and-trends-in-child-under-5-mortality-in-2020. Accessed: 2023 Apr 28.

38. Huber C. Child mortality: Top causes, best solutions. World Vision. 2016. https://www.worldvision.org/health-news-stories/child-mortality-causes-solutions. Accessed: 2023 Apr 28.

39. Ogbuanya AU, Nnadozie UU, Enemuo VC, Ewah RL, Boladuro EO, Owusi OM. Perioperative Mortality Among Surgical Patients in a Low-Resource Setting: A Multi-Center Study at District Hospitals in Southeast Nigeria. Niger J Clin Pract. 2022;25(7):1004.

40. Ouro-Bang’na Maman AF, Agbétra N, Egbohou P, Sama H, Chobli M. Morbidité–mortalité périopératoire dans un pays en développement: expérience du CHU de Lomé (Togo). Ann Fr Anesth Réanimation. 2008;27(12):1030–3.

41. Yunusa A, Cabral C, Anderson E. The impact of the Covid-19 pandemic on the uptake of routine maternal and infant vaccines globally: A systematic review. PLOS Glob Public Health. 2022;2(10):e0000628.

42. PLAN STRATEGIQUE NATIONAL DE LUTTE CONTRE LE PALUDISME AU CAMEROUN 2019-2023. MINSANTE, Yaoundé. 2018.https://lnsp-cam.org/wp-content/uploads/2021/10/PSNLP-2019-2023-CONSOLIDE-TRANSMIS.pdf. Accessed:2023 Apr 30.

